# What contraceptive side effects are women told about during counseling? Evidence from PMA Ethiopia 2019 cross-sectional survey

**DOI:** 10.1101/2023.06.06.23291058

**Authors:** Linnea A. Zimmerman, Isabella Sarria, Munir Kassa, Celia Karp, Assefa Seme, Solomon Shiferaw

**Affiliations:** Department of Population, Family, and Reproductive Health, Johns Hopkins Bloomberg School of Public Health, Baltimore MD, USA; Federal Ministry of Health, Ethiopia, Addis Ababa, Ethiopia; School of Public Health, Addis Ababa University, Addis Ababa, Ethiopia

**Author notes:** Corresponding author: Linnea A. Zimmerman, Johns Hopkins Bloomberg School of Public Health, Department of Population, Family and Reproductive Health, 615 N Wolfe St E4531Batlimore, MD 21231.

## Abstract

**Background:** Despite widespread concerns about contraceptive side effects, few studies explore the specific side effects women are told about during contraceptive counseling. This study explored patterns of counseling on specific side effects by user characteristics in Ethiopia.

**Methods:** Data were collected from a nationally representative sample of women between October and December 2019. Analyses were restricted to 2,039 current users of hormonal contraception or the non-hormonal IUD. Descriptive analyses identified the types and number of side effects, across all methods and by the injectable, implant, and pill. Multinomial regression identified factors associated with receipt of counseling on bleeding changes only, non-bleeding changes only, or both, relative to no counseling on side effects, adjusting for method type, source, and socio-economic characteristics.

**Results:** Less than 10% of users were told of at least one bleeding and non-bleeding side effect. Relative to implant users, injectable and other method users were less likely to be told about bleeding changes only (aRRR: 0.65, 95% CI: 0.46-0.93 and aRRR: 0.31, 95% CI: 0.16-0.61, respectively) and users of other methods were less likely to be told about both a bleeding and non-bleeding change (aRRR: 0.43, 95% CI: 0.19-0.93). Users who received their method from a non-public source were less likely to receive counseling on any kind of side effect and nulliparous women were less likely to be told about both kinds of side effects.

**Conclusion:** There is need to improve counseling in the private sector. Research to understand provider barriers to counseling on side effects is needed.

## Background

The provision of unbiased, scientifically accurate information supports the rights of women to make informed decisions about their reproductive health, including their choice of contraceptive method ^1,2^. Counseling on potential side effects, particularly for hormonal contraceptive methods and the copper IUD, which can induce a range of documented side effects ^3^, is a critical component of counseling. Fear of side effects or health concerns is among the most common reasons provided for not using a method, despite wanting to delay or prevent pregnancy ^4–6^, and the most common reason given for contraceptive discontinuation among women at risk of unintended pregnancy ^7–9^. Counseling that includes information on potential side effects is effective in increasing the likelihood of method adoption and continuation and in decreasing discontinuation rates ^10–12^. Conversely, the provision of inaccurate, incomplete, or biased information can increase dissatisfaction and discontinuation ^13^, and most importantly, is a violation of women’s right to accurate information, limiting their reproductive autonomy ^2,14^.

Recognizing the importance of informing women about potential side effects, recent measurement efforts have prioritized assessing counseling on side effects. Specifically, items measuring the receipt of this information were included in the Method Information Index (MII), a three-question measure evaluating whether a woman was told about 1) methods other than the method she chose, 2) side effects, and 3) what to do if she experienced side effects ^15^. Higher scores on the MII have been shown to be associated with contraceptive behavior; women who reported being informed about all aspects of the MII were 80% less likely to discontinue, while those informed about any one aspect of the MII were 68% less likely to discontinue contraceptive use ^16^. Given its ease in measurement and interpretation, in addition to its association with key contraceptive behaviors ^17^, the MII has been widely used as a proxy to monitor, evaluate, and improve the quality of contraceptive services ^18–21^.

Despite the utility of the MII, however, evidence gaps remain related to the content and comprehensiveness of counseling on side effects. Identifying the side effects on which women are counseled has important implications for program design and intervention. Chang and colleagues asked women in Pakistan and Uganda to report specific side effects on which they were counseled and adjusted the MII to reflect whether these side effects were consistent with their chosen method. When adjusted for method-specific consistency, MII estimates were up to 50% lower in Pakistan and 30% lower in Uganda, indicating that women were frequently counseled on side effects that were not associated with their chosen method ^22^, an important programmatic finding. Similarly, while women frequently report contraceptive-induced menstrual bleeding changes as a major concern ^23^, recent population-based surveys have identified a wide-range of side effects, including some such as headaches and nausea, that are reported as, or more, frequently than bleeding changes among hormonal contraceptive and copper IUD users ^24,25^. Women also frequently report concerns of side effects that are not clinically documented, particularly fears of contraceptive-induced infertility or sub-fecundity ^26–29^, which frequently arises in qualitative research as a primary reason to not use hormonal contraceptive methods ^26,27,30,31^. Understanding the specific side effects on which women are counseled, whether clinically validated or not, is thus critical to design counseling tools and training programs that align with side effects women are likely to experience and/or that are of particular concern, such as infertility.

Such efforts can be further improved by identifying whether the comprehensiveness of side effect counseling varies across women. Recent evidence suggests that women who use long-acting methods are more likely to receive all components of the MII ^19,20^, indicating that method choice may influence the comprehensiveness of counseling services (or vice versa). Similarly, evidence across 25 countries found that the MII was generally positively associated with wealth and education and was higher in urban relative to rural areas and among women who received services from public, relative to private, providers^20^. Additionally, provider-held concerns about contraceptive-induced infertility and other safety concerns may influence counseling for young, unmarried, and nulliparous women ^32–34^. Evidence thus suggests that women of more privileged backgrounds and those who are older and parous may be more likely to receive comprehensive information on side effects, but this has not been documented.

The purpose of this study is thus to explore patterns of specific side effects on which hormonal contraceptive or copper IUD users are counseled, using nationally representative cross-sectional data from Ethiopia. Our objectives are first to describe overall levels of counseling on specific side effects and then to explore patterns in receipt of counseling by user characteristics; specifically, we examine whether receipt of counseling on bleeding changes, non-bleeding changes, or receipt of both varies by sociodemographic characteristics, accounting for method type and source.

## Methods

### Study Setting

Ethiopia is a low-income country in sub-Saharan Africa. Modern contraceptive use has increased significantly, from approximately 8% of married women in 2000 ^35^ to 35.8% in 2019 ^36^. The method mix is largely dominated by the injectable (55% of modern contraception users), but use of the contraceptive implant has grown in recent years, from 16% of the method mix in 2014 to 32% in 2019 ^36^. Despite increases in use, recent evidence from Ejigu and colleagues found that quality of counseling services has declined recently, from 39% of modern contraceptive users receiving all three components of the MII in 2015 to 12% in 2019 ^21^. Concerns regarding side effects are consistently reported as a main contributor to discontinuation ^37–39^, and discontinuation rates within one year remain high, exceeding 40% ^37^.

### Data Source

Data came from the PMA Ethiopia 2019 nationally-representative, cross-sectional household survey of women ages 15-49. PMA Ethiopia is a survey platform implemented in collaboration between Addis Ababa University, the Federal Ministry of Health of Ethiopia, and Johns Hopkins University. Multistage sampling using probability proportional to size within region and urban/rural strata was used to select 265 enumeration areas (EAs). Between October and December 2019, all households within each EA were listed and 35 households were randomly selected. All women aged 15-49 who were either usual members of the household or who slept in the household the night before were eligible for the cross-sectional survey. After being informed of study procedures, women provided verbal consent and were interviewed by a previously trained female interviewer. A total of 8,837 women were interviewed. More information about the study and design are available from the study protocol ^40^. PMA Ethiopia received ethical approval from Addis Ababa University, College of Health Sciences (Ref: AAUMF 01–008) and the Johns Hopkins University Bloomberg School of Public Health Institutional Review Board (FWA00000287). Personally identifiable information collected during data collection were available to the data management team and PIs during data collection, however, all identifiable information was deleted from all datasets prior to data being available for analysis.

### Measures

Our outcome measures explored the specific side effects, number of side effects, and types of side effects on which women were counseled. We identified the specific side effects using the question, *“According to the provider, what are the possible side effects or problems related to use of [current method]”,* where “current method” indicates the current method used by the woman. Answer options included known side effects, such as changes to menstrual bleeding, in addition to side effects that are reported by women, but which are not clinically validated, such as impacts on fertility ^24–26^. Answers were coded based on spontaneous response and were not read aloud. After first exploring the list of all side effects, we evaluate the number of side effects on which women reported being counseled. Finally, we construct a four-category variable exploring whether women were counseled on contraceptive-induced bleeding changes only, non-bleeding changes, both, or neither. Table 1 indicates how side effects were grouped. For regression analyses, described further below, we included socio-demographic characteristics identified from previous literature related to receiving more comprehensive counseling ^19–21^, specifically age (categorical in five year age groups), parity (categorized as 0, 1-2, 3+ births), highest level of schooling attended (none, primary, secondary and above), residence (urban versus rural), and wealth quintile. Recent evidence has also highlighted critical differences in the MII across regions and by method source in Ethiopia ^21^ and we thus include region (Oromiya, Amhara, Addis, Southern Nations, Nationalities, and Peoples Region (SNNP-R)^1^, Tigray^2^, and all other regions) and method source (hospital or health center, health post or HEW, private facilities and pharmacies).

**Table 1:** Percentage of women who were told about 0, 1, 2, 3, or 4 plus sideeffects among current hormonal contraceptive or copper IUD users

|  |  |  |  |  |
| --- | --- | --- | --- | --- |
|  | All users | Implant Users | Injectable Users | Pill Users |
|  | % | % | % | % |
| 0 | 73.0 | 64.8 | 76.1 | 82.8 |
| 1 | 10.5 | 12.9 | 9.5 | 8.7 |
| 2 | 7.0 | 8.0 | 6.6 | 6.5 |
| 3 | 4.8 | 8.0 | 3.5 | 1.0 |
| 4+ | 4.8 | 6.2 | 4.4 | 1.0 |

### Analyses

#### Analytic Sample and Analysis

Our analytic sample was restricted to women who slept in the household the night before, completed the female questionnaire, and were current users of a hormonal modern method of contraception (implant, injectable, pill, or emergency contraception) or the copper IUD (n=2,039). Descriptive statistics of frequencies and percentages were used to explore patterns of specific side effects and number of side effects on which women were counseled, among all hormonal contraception and copper IUD users and stratified by the three most-commonly used methods (implant, injectable, pill). Sample size limitations prevented stratification by EC or the copper IUD. We then used unadjusted and adjusted multinomial logistic regression models to explore the relationship between receipt of type of side effect counseling and women’s sociodemographic characteristics, using women who received no side effect counseling as the reference. Due to high correlation between age and parity (ρ=0.70), and wealth and residence (ρ=0.73), we do not include age or wealth in the adjusted regression model. Additionally, as marriage is almost universal in the sample (Appendix Table 1), we did not adjust for marital status. Due to sample size limitations, we combine pill, EC, and copper IUD users into one “other” method type. Analyses accounted for complex survey design through application of weights and adjustment for clustering and were conducted using Stata SE 16.1 ^41^.

## Results

### Sociodemographic characteristics of users are shown in Appendix Table 1

Table 1 below shows the number of side effects on which women were counseled, across all users and by method. Across all users, almost three in four women report not receiving counseling on any side effects. Among women who do receive counseling, most received counseling on only one side effect. A higher percentage of implant users reported receiving any counseling on side effects (35.2%) and on more than one side effect (20.2%) than injectable (23.9% and 14.5%, respectively) or pill users (17.2% and 8.5%, respectively). Pill users received the least amount of side effect counseling, relative to implant and injectable users.

Table 2 shows the percentage of users who reported being told about each of the side effects, shown among all current hormonal users and by implant, injectable, and pill users. The most common side effects on which women were counseled were related to menstrual bleeding changes. Fewer than ten percent of women reported receiving counseling about any other potential side effects.

**Table 2:** Percentage of current hormonal contraception or copper IUD users who received counseling on each side-effect

|  |  |  |  |  |
| --- | --- | --- | --- | --- |
|  | All current users | Implant users | Injectable users | Pill users |
|  | % | % | % | % |
| **N** | **2039** | **675** | **1,114** | **158** |
| **Bleeding changes** |  |  |  |  |
| Less or No Bleeding | 12.7 | 15.7 | 12.5 | 1.7 |
| More Bleeding | 10.7 | 15.3 | 8.8 | 4.2 |
| Irregular Bleeding | 8.8 | 12.8 | 7.2 | 2.4 |
| Spotting | 3.7 | 5.6 | 2.7 | 0.9 |
| General Bleeding (non-specific) | 2.6 | 3.5 | 2.3 | 0.9 |
| **Non-bleeding changes** |  |  |  |  |
| Abdominal Pain | 0.9 | 0.8 | 1.0 | 0.6 |
| Gain Weight | 3.7 | 4.6 | 3.5 | 1.7 |
| Lose Weight | 3.7 | 4.4 | 3.7 | 2.3 |
| Acne | 1.4 | 1.0 | 1.6 | 1.8 |
| Headache | 5.8 | 9.6 | 3.8 | 4.3 |
| Infection | 0.4 | 0.7 | 0.3 | 0.6 |
| Nausea | 1.0 | 1.5 | 0.6 | 1.6 |
| Menstrual Cramp | 1.0 | 0.1 | 1.4 | 2.4 |
| Lower sex drive | 0.3 | 0.3 | 0.3 | 0.0 |
| Vaginal dryness | 0.2 | 0.1 | 0.2 | 0.0 |
| Infertility/Sterility | 1.0 | 0.1 | 1.5 | 0.8 |
| Delayed Fertility | 1.1 | 0.2 | 1.7 | 1.3 |
| Method Lost | 0.3 | 0.7 | 0.1 | 0.0 |
| Weakness | 2.1 | 3.4 | 1.4 | 0.0 |

Table 3 below shows the distribution of types of side effects on which women were counseled. About 20% of all users were counseled on at least one contraceptive-induced menstrual bleeding side effect and about 15% were counseled on a non-bleeding side effect. A higher percentage of implant users (12.9%) reported being told about at least one bleeding and one non-bleeding side effect, but less than five percent of either injectable or pill users reported receiving counseling on both types of side effects.

**Table 3:** Percentage of women who were told about different kinds of side effects, by method type

|  |  |  |  |  |
| --- | --- | --- | --- | --- |
|  | All users | Implant Users | Injectable Users | Pill Users |
|  | % | % | % | % |
| No side effect | 73.0 | 64.8 | 76.1 | 82.8 |
| Bleeding only | 11.7 | 16.2 | 10.1 | 3.5 |
| Non-bleeding only | 6.0 | 6.9 | 5.2 | 10.1 |
| Both | 9.4 | 12.9 | 3.6 | 3.6 |

Unadjusted results assessing the relationship between method choice, selected sociodemographic and environmental factors, and receipt of type of side effect counseling are shown in Table 4 below. Injectable and “other” method users (pill, EC, and copper IUD) were significantly less likely to be told only about a bleeding change (RRR: 0.53, 95% CI: 0.37-0.75 and RRR: 0.27, 95% CI: 0.14-0.52, respectively) and significantly less likely to be told about both a bleeding change and a non-bleeding side effect (RRR: 0.62, 95% CI: 0.40-0.94 and RRR: 0.33, 95% CI: 0.16-0.68, respectively). Parous women were significantly more likely to receive counseling on at least one bleeding and non-bleeding side effect than nulliparous women. Women who received their method from a private source were significantly less likely to receive counseling on bleeding changes only or on both bleeding changes and non-bleeding side effects (RRR: 0.42, 95% CI: 0.26-0.69 and RRR: 0.23, 95% CI: 0.12-0.44, respectively) while women who received services from a health post or HEW were significantly less likely to receive counseling on at least one of each type of side effect than women who received services from a hospital or health center (RRR: 0.59, 95% CI: 0.36-0.98). Region, wealth, and education appeared to be largely unrelated to the type of side effect on which women were counseled, with some exceptions.

**Table 4:** Unadjusted multinomial logistic regression results. PMA Ethiopia 2019 cross-sectional survey

|  |  | CIMBC only |  |  | non-CIMBC only |  |  | Both |  |  |
| --- | --- | --- | --- | --- | --- | --- | --- | --- | --- | --- |
|  |  | RRR | 95% CI |  | RRR | 95% CI |  | RRR | 95% CI |  |
| **Method (ref: implant)** |  |  |  |  |  |  |  |  |  |  |
|  | Injectable | 0.53 | 0.37 | 0.75 | 0.63 | 0.38 | 1.06 | 0.62 | 0.40 | 0.94 |
|  | Other | 0.27 | 0.14 | 0.52 | 0.82 | 0.45 | 1.50 | 0.33 | 0.16 | 0.68 |
| **Age (ref: 15-19)** |  |  |  |  |  |  |  |  |  |  |
|  | 20-24 | 0.75 | 0.35 | 1.64 | 0.80 | 0.33 | 1.93 | 1.79 | 0.68 | 4.72 |
|  | 25-29 | 1.12 | 0.58 | 2.16 | 1.35 | 0.59 | 3.08 | 2.86 | 1.11 | 7.38 |
|  | 30-34 | 1.17 | 0.57 | 2.43 | 0.66 | 0.26 | 1.67 | 2.19 | 0.85 | 5.64 |
|  | 35-39 | 1.04 | 0.57 | 1.90 | 0.93 | 0.36 | 2.39 | 1.68 | 0.64 | 4.41 |
|  | 40-44 | 0.78 | 0.32 | 1.93 | 1.02 | 0.38 | 2.70 | 2.51 | 0.82 | 7.66 |
|  | 45-49 | 0.81 | 0.27 | 2.47 | 0.57 | 0.08 | 3.97 | 2.08 | 0.48 | 9.01 |
| **Parity (ref: 0)** |  |  |  |  |  |  |  |  |  |  |
|  | 1-2 | 1.84 | 0.92 | 3.67 | 1.10 | 0.54 | 2.23 | 3.05 | 1.49 | 6.23 |
|  | 3+ | 1.69 | 0.84 | 3.42 | 1.11 | 0.53 | 2.33 | 2.47 | 1.16 | 5.24 |
| **Education (ref: None)** |  |  |  |  |  |  |  |  |  |  |
|  | Primary | 1.13 | 0.78 | 1.64 | 1.19 | 0.72 | 1.98 | 1.05 | 0.69 | 1.58 |
|  | Secondary | 1.38 | 0.93 | 2.05 | 1.63 | 0.90 | 2.95 | 1.73 | 1.06 | 2.83 |
| **Residence (ref: rural)** |  |  |  |  |  |  |  |  |  |  |
|  | Urban | 1.09 | 0.74 | 1.61 | 1.53 | 0.92 | 2.56 | 1.03 | 0.67 | 1.59 |
| **Region (ref: Oromiya)** |  |  |  |  |  |  |  |  |  |  |
|  | Tigray | 0.38 | 0.23 | 0.64 | 0.94 | 0.47 | 1.89 | 0.81 | 0.47 | 1.39 |
|  | Amhara | 0.95 | 0.56 | 1.62 | 1.72 | 0.83 | 3.56 | 1.15 | 0.65 | 2.03 |
|  | Addis | 0.62 | 0.33 | 1.16 | 0.76 | 0.35 | 1.68 | 1.14 | 0.60 | 2.16 |
|  | SNNP | 0.74 | 0.37 | 1.48 | 1.11 | 0.55 | 2.24 | 0.78 | 0.34 | 1.77 |
|  | Other | 0.83 | 0.51 | 1.36 | 2.39 | 1.22 | 4.68 | 1.63 | 0.76 | 3.50 |
| **Wealth (ref: Lowest)** |  |  |  |  |  |  |  |  |  |  |
|  | Lower | 0.79 | 0.40 | 1.55 | 0.38 | 0.15 | 0.98 | 0.87 | 0.42 | 1.80 |
|  | Middle | 1.29 | 0.60 | 2.78 | 0.53 | 0.20 | 1.40 | 1.22 | 0.63 | 2.37 |
|  | Higher | 1.27 | 0.67 | 2.40 | 0.94 | 0.42 | 2.12 | 0.92 | 0.40 | 2.10 |
|  | Highest | 1.35 | 0.74 | 2.44 | 1.03 | 0.47 | 2.24 | 1.27 | 0.62 | 2.61 |
| **Source of method (ref: Hospital/Health Center)** |  |  |  |  |  |  |  |  |  |  |
|  | Health Post/HEW | 0.76 | 0.49 | 1.17 | 0.73 | 0.39 | 1.37 | 0.59 | 0.36 | 0.98 |
|  | Non-public | 0.42 | 0.26 | 0.69 | 0.56 | 0.30 | 1.02 | 0.23 | 0.12 | 0.44 |

Adjusted results are shown in Table 5. Results were largely consistent after adjustment. Injectable and other method users were significantly less likely than implant users to be told only about only a bleeding side effect and other method users were significantly less likely to be told about both a bleeding and non-bleeding side effect. Women of higher parity remained significantly more likely to be told about both a bleeding and non-bleeding side effect. Women with secondary education and higher were more likely to receive counseling on bleeding changes only or on both bleeding-and non-bleeding changes.

**Table 5:**
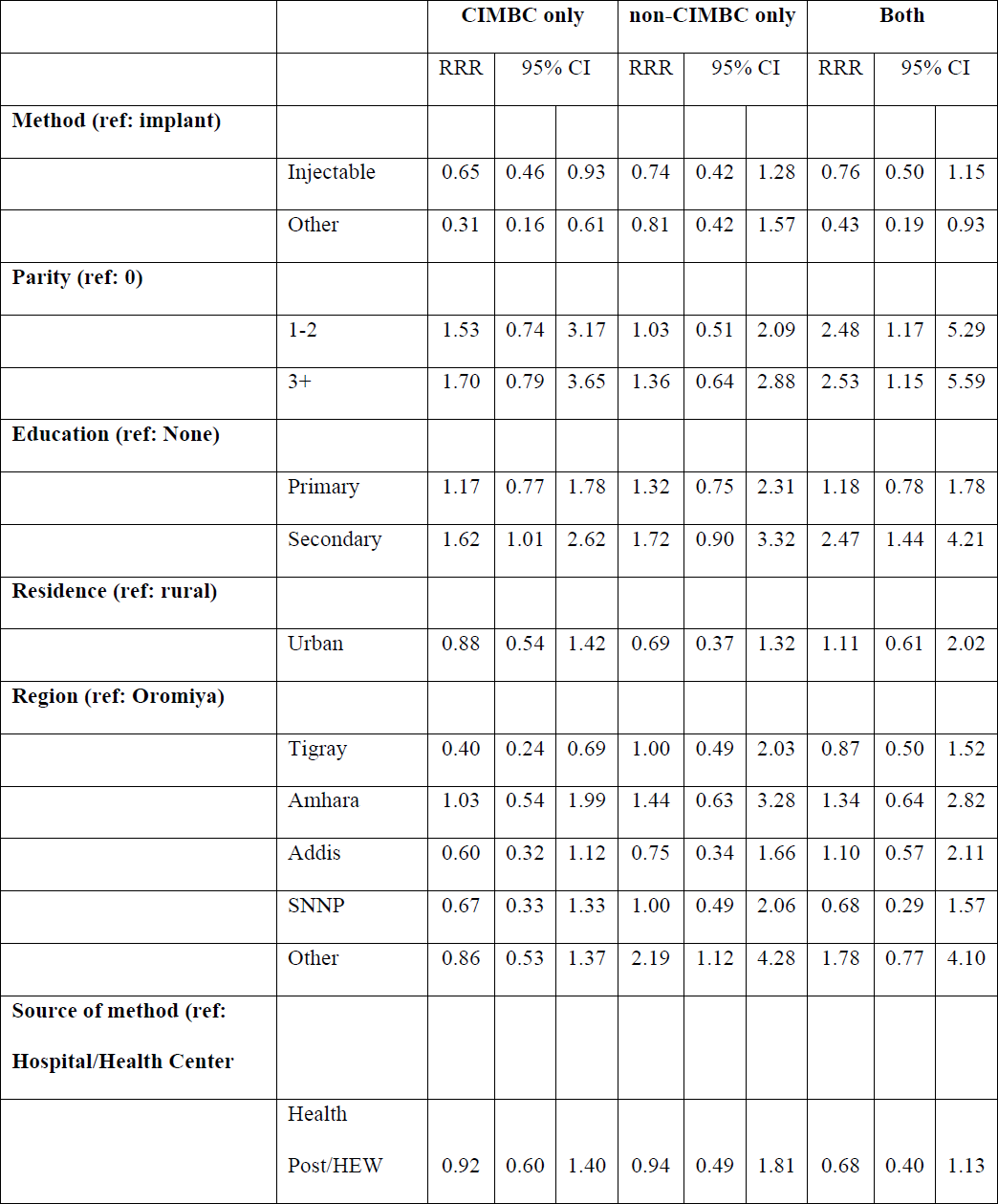
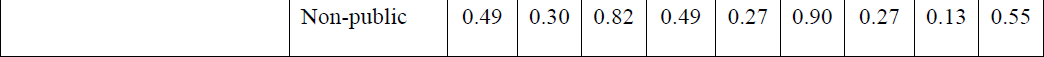
Adjusted multinomial logistic regression results. PMA Ethiopia 2019 cross-sectional survey

## Discussion

We find that most contraceptive users in Ethiopia do not receive sufficient counseling, as approximately three in four users reported they were either not counseled on any side effects or received counseling on only one side effect. Women were most likely to receive counseling on contraceptive-induced menstrual bleeding changes, if they received any counseling at all, and few women received counseling on any non-bleeding changes. Our findings highlight the ongoing need for improvements in access to high-quality, comprehensive contraceptive services.

Despite current and recent contraceptive users in Ethiopia experiencing a wide range of side effects ^24^, fewer than 20% of women reported being counseled on at least two side effects, and the majority of women who received counseling only received counseling on contraceptive-induced menstrual bleeding changes. While concerns about bleeding changes do feature heavily in women’s worries related to contraceptive side effects ^23^, given the range of potential side effects associated with hormonal contraception and the non-hormonal IUD, this limited counseling does not represent sufficient information exchange between providers and clients. Evidence suggests that women learn about side effects, both clinically validated and not, from a range of sources ^6,31,42^, but largely trust providers to deliver scientifically sound and accurate information ^13,43^. If women are already aware of and concerned about potential side effects, receiving counseling from providers on the full range of potential side effects is an important opportunity to address concerns and dispel myths. Incorporation of structured tools such as the NORMAL ^44^ tool or the Balanced Counseling Strategy ^45^ could help address some of these counseling gaps and help women make informed decisions about contraception. The Federal Ministry of Health of Ethiopia recently introduced a strategy to improve counseling through the use of structured counseling tools in public health facilities, but implementation was at the onset at the time of this publication.

Concerns related to contraceptive-induced fertility impairment may be particularly relevant to address, as these concerns, at least qualitatively, remain a major reason for contraceptive non-use ^26,27^. Approximately 1% of users reported that they were counseled about infertility or delayed return to fertility. A significant weakness in our study is that we cannot distinguish whether providers were reinforcing or dispelling these concerns, however, our finding that so few women reported being counseled on this topic reinforces that women are unlikely to have this concern addressed by a provider. Additionally, we found that women who were more educated and those who had a greater number of children were more likely to receive counseling on at least one potential bleeding change and one non-bleeding change related to their contraceptive method. While our study is unable to explain what specific behaviors would influence why nulliparous women would receive less comprehensive counseling, fears related to contraception may be particularly relevant to address among younger and nulliparous women as they tend to have the lowest rates of contraceptive use among women who wish to prevent pregnancy ^46,47^. Provider bias towards specific methods based on patient characteristics, such as marital status and parity, may influence counseling, as has been documented elsewhere ^13,14,34,48^. Additional research among providers is necessary to understand how user characteristics influence providers’ counseling practices on side effects.

Finally, receipt of more comprehensive counseling was influenced by method choice and source of method. On the whole, implant users were the most likely to receive counseling on side effects and pill users were the least likely. While side effect profiles differ by method and thus, counseling should also differ, all hormonal methods have the potential for both bleeding and non-bleeding changes. As the implant requires clinical intervention to stop use, it is potentially positive that implant users received relatively more counseling on side effects, however, all women, regardless of the method they select, should be comprehensively counseled in order to make an informed choice. As with directive counseling based on patient characteristics, differences in counseling by method may be explained by provider concerns about safety or preferences for specific methods ^34^. This finding further reinforces the need for research with providers to understand how knowledge of and concerns about side effects influence counseling.

These method-specific differences in counseling experiences may in part be explained by method source. For example, pill users may be more likely to source their method from a range of sources and less likely to rely on trained providers. This relationship between method type and counseling remained, however, even after adjusting for method source. Women who received their methods outside of the public health sector had significantly lower relative risk of receiving counseling on any side effects. The majority of women in Ethiopia access their methods through the public health sector ^36^, but efforts are underway to expand the private sector in Ethiopia through such projects as the Ethiopia Private Health Sector Project ^49,50^. Evidence suggests that private facilities are less likely to have trained FP providers or clear guidelines and protocols for FP provision ^51^. Thus, expansion of the private sector must be done with quality in mind, ensuring that private providers, particularly drug shop owners and pharmacists, are trained to provide comprehensive counseling.

Our study is not without limitations. Our survey question about contraceptive side effects did not distinguish what information was provided about each side effect or confirm if scientifically accurate information was shared. Such nuance is likely not possible in a population-based survey and requires alternative data collection strategies, such as observation of counseling sessions. Additionally, we collected information retroactively; recall of specific information is likely to fade over time and introduce bias. If so, we may be underestimating the percentage of women who received more comprehensive counseling. Similarly, we collect information only among current users of contraception. Women who discontinued their method prior to the survey or who received counseling but chose not to adopt a method may have received different counseling than women who continued, which is not reflected in our findings due to data limitations. Finally, data were collected in Tigray in 2019 prior to the onset of widespread civil conflict and are not able to be generalized to the current situation. Despite these limitations, we believe that the use a high-quality, nationally representative data source that incorporated detailed information on side effect counseling provides clear evidence of gaps in counseling that must be addressed to enhance informed decision-making and improve reproductive health outcomes.

## Conclusions

Women in Ethiopia do not receive sufficient information on side effects during contraceptive counseling, particularly related to side effects outside of menstrual bleeding changes. Private health facilities appear to be of particular concern. Efforts to improve counseling, such as structured counseling tools, should be incorporated into the private sector. Additionally, research with providers is necessary to understand and address biases towards specific methods and counseling messages.

## Data Availability

Data are available from https://www.pmadata.org/data/request-access-datasets.

## Declarations

### Acknowledgements

We thank the PMA Ethiopia team for their dedication in conducting the PMA Ethiopia surveys and the respondents for sharing their time and experiences.

**Appendix Table 1:** Sample characteristics of users of hormonal contraception and the IUD; PMA Ethiopia Cross-section 2019

|  |  |  |  |
| --- | --- | --- | --- |
|  |  | Total |  |
|  |  | n<br>(unweighted) | %<br>(weighted) |
| Method |  |  |  |
|  | Implant | 675 | 33.2 |
|  | Injectable | 1114 | 57.4 |
|  | Pill | 158 | 6.5 |
|  | Other | 73 | 2.9 |
| Source of Method |  |  |  |
|  | Hospital/health center | 1103 | 53.4 |
|  | Health post/HEW | 464 | 27.0 |
|  | Non-public | 472 | 19.6 |
| Age |  |  |  |
|  | 15-19 | 160 | 9.0 |
|  | 20-24 | 439 | 21.4 |
|  | 25-29 | 578 | 27.4 |
|  | 30-34 | 372 | 16.8 |
|  | 35-39 | 288 | 14.3 |
|  | 40-44 | 160 | 9.1 |
|  | 45-49 | 42 | 2.2 |
| Marital Status |  |  |  |
|  | Married | 202 | 8.4 |
|  | Unmarried | 1837 | 91.6 |
| Parity |  |  |  |
|  | 0 | 238 | 10.8 |
|  | 1-2 | 945 | 44.2 |
|  | 3+ | 855 | 45.0 |
| Education |  |  |  |
|  | None | 648 | 36.4 |
|  | Primary | 790 | 39.6 |
|  | Secondary + | 601 | 23.9 |
| Residence |  |  |  |
|  | Rural | 1100 | 64.1 |
|  | Urban | 939 | 35.9 |
| Region |  |  |  |
|  | Oromiya | 449 | 38.8 |
|  | Amhara | 451 | 27.7 |
|  | Addis | 206 | 6.4 |
|  | SNNP | 407 | 19.4 |
|  | Tigray | 243 | 5.2 |
|  | Other | 283 | 2.5 |
| Wealth |  |  |  |
|  | Lowest | 241 | 15.0 |
|  | Lower | 305 | 18.3 |
|  | Middle | 360 | 21.3 |
|  | Higher | 421 | 19.9 |
|  | Highest | 712 | 25.5 |

## Footnotes

1 In 2020, a new region, Sidama, was formed from within SNNP-R. Data were collected prior to the creation of this region.

2 In 2020, civil conflict began in Tigray leading to a protracted humanitarian emergency. Data were collected prior to the onset of this conflict and are not likely to represent the current situation.

## References

1. Bruce J. Fundamental Elements of the Quality of Care: A Simple Framework. Studies in Family Planning. 1990;21(2):61. doi:10.2307/1966669

2. Hardee K. Operationalizing Human Rights in Sexual and Reproductive Health and Rights Programming: An Example from a Global Family Planning Partnership. Oxford Research Encyclopedia of Global Public Health. doi:10.1093/acrefore/9780190632366.013.239

3. Hatcher RA, Trussell J, Nelson AL, Cates Jr W, Kowal D, Policar MS. Contraceptive Technology. 20th Revised. Ardent Media, Inc; 2015.

4. Sedgh G, Hussain R. Reasons for Contraceptive Nonuse among Women Having Unmet Need for Contraception in Developing Countries. Studies in Family Planning. 2014;45(2):151–169. doi:10.1111/j.1728-4465.2014.00382.x

5. Schrumpf LA, Stephens MJ, Nsarko NE, et al. Side effect concerns and their impact on women’s uptake of modern family planning methods in rural Ghana: a mixed methods study. BMC Womens Health. 2020;20:57. doi:10.1186/s12905-020-0885-0

6. Diamond-Smith N, Campbell M, Madan S. Misinformation and fear of side-effects of family planning. Culture, Health & Sexuality. 2012;14(4):421–433. doi:10.1080/13691058.2012.664659

7. Ali MM, Cleland JG, Shah IH, World Health Organization. Causes and Consequences of Contraceptive Discontinuation: Evidence from 60 Demographic and Health Surveys. World Health Organization; 2012. Accessed February 7, 2022. https://apps.who.int/iris/handle/10665/75429

8. Bradley SEK, Schwandt HM, Khan S. Levels, Trends, and Reasons for Contraceptive Discontinuation. ICF Macro; 200AD. Accessed March 5, 2021. https://www.dhsprogram.com/pubs/pdf/AS20/AS20.pdf

9. Jain A, Reichenbach L, Ehsan I, Rob U. “Side effects affected my daily activities a lot”: a qualitative exploration of the impact of contraceptive side effects in Bangladesh. Open Access J Contracept. 2017;8:45–52. doi:10.2147/OAJC.S140214

10. Canto De Cetina TE, Canto P, Ordoñez Luna M. Effect of counseling to improve compliance in Mexican women receiving depot-medroxyprogesterone acetate. Contraception. 2001;63(3):143–146. doi:10.1016/S0010-7824(01)00181-0

11. Dehingia N, Dixit A, Averbach S, et al. Family planning counseling and its associations with modern contraceptive use, initiation, and continuation in rural Uttar Pradesh, India. Reprod Health. 2019;16(1):178. doi:10.1186/s12978-019-0844-0

12. Huda FA, Chowdhuri S, Sirajuddin MFR. Importance of Appropriate Counselling in Reducing Early Discontinuation of Norplant in a Northern District of Bangladesh. J Health Popul Nutr. 2014;32(1):142–148.

13. Yirgu R, Wood SN, Karp C, Tsui A, Moreau C. “You better use the safer one… leave this one”: the role of health providers in women’s pursuit of their preferred family planning methods. BMC Women’s Health. 2020;20(1):170. doi:10.1186/s12905-020-01034-1

14. Senderowicz L, Pearson E, Hackett K, et al. ‘I haven’t heard much about other methods’: quality of care and person-centredness in a programme to promote the postpartum intrauterine device in Tanzania. BMJ Global Health. 2021;6(6):e005775. doi:10.1136/bmjgh-2021-005775

15. Jain A, Townshend J, RamaRao S. Proposed Metrics to Measure Quality: Overview. Population Council; 2018. doi:10.31899/rh6.1024

16. Chakraborty NM, Chang K, Bellows B, et al. Association Between the Quality of Contraceptive Counseling and Method Continuation: Findings From a Prospective Cohort Study in Social Franchise Clinics in Pakistan and Uganda. Glob Health Sci Pract. 2019;7(1):87–102. doi:10.9745/GHSP-D-18-00407

17. Tin KN, Maung TM, Win T. Factors that affect the discontinuation of family planning methods in Myanmar: analysis of the 2015–16 Myanmar Demographic and Health Survey. Contracept Reprod Med. 2020;5(1):20. doi:10.1186/s40834-020-00126-5

18. Budiharsana MP, Wahyuningsih W, Heywood P. The use of Method Information Index (MII) to monitor the amount of information given to women users of modern contraceptives in Indonesia: results from an analysis of the 2007, 2012 and 2017 demographic and health surveys. BMC Women’s Health. 2022;22(1):489. doi:10.1186/s12905-022-02094-1

19. Bullington BW, Tumlinson K, Karp C, et al. Do users of long-acting reversible contraceptives receive the same counseling content as other modern method users? A cross-sectional, multi-country analysis of women’s experiences with the Method Information Index in six sub-Saharan African countries. Contracept X. 2022;4:100088. doi:10.1016/j.conx.2022.100088

20. Jain. Examining Progress and Equity in Information Received by Women Using a Modern Method in 25 Developing Countries. International Perspectives on Sexual and Reproductive Health. 2016;42(3):131. doi:10.1363/42e1616

21. Ejigu BA, Seme A, Zimmerman L, Shiferaw S. Trend and determinants of quality of family planning counseling in Ethiopia: Evidence from repeated PMA cross-sectional surveys, (2014–2019). PLOS ONE. 2022;17(5):e0267944. doi:10.1371/journal.pone.0267944

22. Chang KT, Mukanu M, Bellows B, et al. Evaluating Quality of Contraceptive Counseling: An Analysis of the Method Information Index. Studies in Family Planning. 2019;50(1):25–42. doi:10.1111/sifp.12081

23. Polis CB, Hussain R, Berry A. There might be blood: a scoping review on women’s responses to contraceptive-induced menstrual bleeding changes. Reprod Health. 2018;15(1):114. doi:10.1186/s12978-018-0561-0

24. Zimmerman LA, Sarnak DO, Karp C, et al. Measuring experiences and concerns surrounding contraceptive induced side-effects in a nationally representative sample of contraceptive users: Evidence from PMA Ethiopia. Contraception: X. 2022;4:100074. doi:10.1016/j.conx.2022.100074

25. Zimmerman LA, Sarnak DO, Karp C, et al. Association between experience of specific side-effects and contraceptive switching and discontinuation in Uganda: results from a longitudinal study. Reproductive Health. 2021;18(1):239. doi:10.1186/s12978-021-01287-5

26. Sedlander E, Bingenheimer JB, Thiongo M, et al. “They Destroy the Reproductive System”: Exploring the Belief that Modern Contraceptive Use Causes Infertility. Studies in Family Planning. 2018;49(4):345–365. doi:https://doi.org/10.1111/sifp.12076

27. Sedlander E, Bingenheimer JB, Lahiri S, et al. Does the Belief That Contraceptive Use Causes Infertility Actually Affect Use? Findings from a Social Network Study in Kenya. Studies in Family Planning. 2021;52(3):343–359. doi:10.1111/sifp.12157

28. Sedlander E, Yilma H, Emaway D, Rimal RN. If fear of infertility restricts contraception use, what do we know about this fear? An examination in rural Ethiopia. Reproductive Health. 2022;19(1):57. doi:10.1186/s12978-021-01267-9

29. Boivin J, Carrier J, Zulu JM, Edwards D. A rapid scoping review of fear of infertility in Africa. Reproductive Health. 2020;17(1):142. doi:10.1186/s12978-020-00973-0

30. Adofo E, Dun-Dery EJ, Kotoh AM, Dun-Dery F, Avoka JA, Ashinyo ME. Fear of infertility limits contraceptive usage among first-time mothers in Ghana: A cross-sectional study. SAGE Open Medicine. 2021;9:20503121211021256. doi:10.1177/20503121211021256

31. Schwarz J, Dumbaugh M, Bapolisi W, et al. “So that’s why I’m scared of these methods”: Locating contraceptive side effects in embodied life circumstances in Burundi and eastern Democratic Republic of the Congo. Soc Sci Med. 2019;220:264–272. doi:10.1016/j.socscimed.2018.09.030

32. Nalwadda G, Mirembe F, Tumwesigye NM, Byamugisha J, Faxelid E. Constraints and prospects for contraceptive service provision to young people in Uganda: providers’ perspectives. BMC Health Serv Res. 2011;11(1):220. doi:10.1186/1472-6963-11-220

33. Paul M, Näsström SB, Klingberg-Allvin M, Kiggundu C, Larsson EC. Healthcare providers balancing norms and practice: challenges and opportunities in providing contraceptive counselling to young people in Uganda – a qualitative study. Global Health Action. 2016;9(1):30283. doi:10.3402/gha.v9.30283

34. Solo J, Festin M. Provider Bias in Family Planning Services: A Review of Its Meaning and Manifestations. Glob Health Sci Pract. 2019;7(3):371–385. doi:10.9745/GHSP-D-19-00130

35. Central Statistical Agency (CSA), ORC Macro. Ethiopia Demographic and Health Survey 2000. Central Stastical Authority and ORC Macro; 2001. Accessed February 10, 2021. https://dhsprogram.com/pubs/pdf/FR328/FR328.pdf

36. PMA Ethiopia. PMA Ethiopia: Results from the 2019 Surveys. Addis Ababa University, School of Public Health and Johns Hopkins Bloomberg School of Public Health; 2020. Accessed January 13, 2023. https://www.pmadata.org/sites/default/files/data_product_results/Ethiopia_2019_Survey_Re sults_Brief.pdf

37. Tsui AO, Brown W, Li Q. Contraceptive Practice in Sub-Saharan Africa. Popul Dev Rev. 2017;43(Suppl Suppl 1):166-191. doi:10.1111/padr.12051

38. Alvergne A, Stevens R, Gurmu E. Side effects and the need for secrecy: characterising discontinuation of modern contraception and its causes in Ethiopia using mixed methods. Contracept Reprod Med. 2017;2(1):24. doi:10.1186/s40834-017-0052-7

39. Abebe M, Mersha A, Degefa N, Gebremeskel F, Kefelew E, Molla W. Determinants of induced abortion among women received maternal health care services in public hospitals of Arba Minch and Wolayita Sodo town, southern Ethiopia: unmatched case–control study. BMC Women’s Health. 2022;22(1):107. doi:10.1186/s12905-022-01695-0

40. Zimmerman L, Desta S, Yihdego M, et al. Protocol for PMA-Ethiopia: A new data source for cross-sectional and longitudinal data of reproductive, maternal, and newborn health. Gates Open Res. 2020;4. doi:10.12688/gatesopenres.13161.1

41. StataCorp. Stata Statistical Software: Release 16. Published online 2019.

42. Chebet JJ, McMahon SA, Greenspan JA, et al. “Every method seems to have its problems”-Perspectives on side effects of hormonal contraceptives in Morogoro Region, Tanzania. BMC Women’s Health. 2015;15(1):97. doi:10.1186/s12905-015-0255-5

43. Keesara SR, Juma PA, Harper CC. Why do women choose private over public facilities for family planning services? A qualitative study of post-partum women in an informal urban settlement in Kenya. BMC Health Serv Res. 2015;15(1):335. doi:10.1186/s12913-015-0997-7

44. Rademacher KH, Sergison J, Glish L, et al. Menstrual Bleeding Changes Are NORMAL: Proposed Counseling Tool to Address Common Reasons for Non-Use and Discontinuation of Contraception. Glob Health Sci Pract. 2018;6(3):603–610. doi:10.9745/GHSP-D-18-00093

45. Population Council. The Balanced Counseling Strategy Plus: A Toolkit for Family Planning Service Providers Working in High HIV/STI Prevalence Settings. Population Council; 2015.

46. de Vargas Nunes Coll C, Ewerling F, Hellwig F, de Barros AJD. Contraception in adolescence: the influence of parity and marital status on contraceptive use in 73 low-and middle-income countries. Reproductive Health. 2019;16(1):21. doi:10.1186/s12978-019-0686-9

47. Behrman JA, Wright KQ, Grant MJ, Soler-Hampejsek E. Trends in Modern Contraceptive Use among Young Adult Women in sub-Saharan Africa 1990 to 2014. Studies in Family Planning. 2018;49(4):319–344. doi:10.1111/sifp.12075

48. Daniele MAS, Cleland J, Benova L, Ali M. Provider and lay perspectives on intra-uterine contraception: a global review. Reproductive Health. 2017;14(1):119. doi:10.1186/s12978-017-0380-8

49. Ministry of Health - Ethiopia, Global Financing Facility, World Bank Group. Ethiopia Health Private Sector Assessment.; 2019. Accessed October 25, 2022. https://www.globalfinancingfacility.org/sites/gff_new/files/documents/Ethiopia-health-private-sector-assessment.pdf

50. The Ethiopia Private Health Sector Project | Abt Associates. Accessed March 21, 2023. https://www.abtassociates.com/projects/the-ethiopia-private-health-sector-project

51. Tessema GA, Mahmood MA, Gomersall JS, et al. Structural Quality of Services and Use of Family Planning Services in Primary Health Care Facilities in Ethiopia. How Do Public and Private Facilities Compare? International Journal of Environmental Research and Public Health. 2020;17(12):4201. doi:10.3390/ijerph17124201

